# ICU and ventilator mortality among critically ill adults with COVID-19

**DOI:** 10.1101/2020.04.23.20076737

**Authors:** Sara C. Auld, Mark Caridi-Scheible, James M. Blum, Chad Robichaux, Colleen Kraft, Jesse T. Jacob, Craig S. Jabaley, David Carpenter, Roberta Kaplow, Alfonso C. Hernandez-Romieu, Max W. Adelman, Greg S. Martin, Craig M. Coopersmith, David J. Murphy, the Emory COVID-19 Quality and Clinical Research Collaborative

**Affiliations:** Emory Critical Care Center (ECCC); Division of Pulmonary, Allergy, Critical Care and Sleep Medicine, Department of Medicine, Emory University School of Medicine; Department of Epidemiology, Emory University Rollins School of Public Health; Department of Anesthesiology, Emory University School of Medicine; Department of Biomedical Informatics, Emory University School of Medicine; Georgia Clinical and Translational Science Alliance (CTSA); Division of Infectious Diseases, Department of Medicine, Emory University School of Medicine; Department of Pathology, Emory University School of Medicine; Emory University Hospital, Emory Healthcare; Department of Surgery, Emory University School of Medicine; Office of Quality and Risk, Emory Healthcare

## Abstract

We report preliminary data from a cohort of adults admitted to COVID-designated intensive care units from March 6 through April 17, 2020 across an academic healthcare system. Among 217 critically ill patients, mortality for those who required mechanical ventilation was 29.7% (49/165), with 8.5% (14/165) of patients still on the ventilator at the time of this report. Overall mortality to date in this critically ill cohort is 25.8% (56/217), and 40.1% (87/217) patients have survived to hospital discharge. Despite multiple reports of mortality rates exceeding 50% among critically ill adults with COVID-19, particularly among those requiring mechanical ventilation, our early experience indicates that many patients survive their critical illness.

## Introduction

COVID-19 has become one of the leading causes of death worldwide. It is estimated that 15–20% of cases require hospitalization and 3–5% require critical care. While experience with COVID-19 continues to grow, reported mortality rates range from 50–97% in those requiring mechanical ventilation.^1-6^ These are significantly higher than the published mortality rates ranging from 35–46% for patients intubated with H1N1 influenza pneumonia and other causes of acute respiratory distress syndrome (ARDS).^7-10^

These high mortality rates have raised concerns as to whether invasive mechanical ventilation should be avoided in the context of COVID-19.^11-14^ To help address the growing concern that critical illness, and specifically mechanical ventilation, are associated with a high risk of death, we conducted a retrospective cohort study of critically ill patients with COVID-19 across our academic health system.

## Methods

This is an observational cohort study of all patients with COVID-19 admitted to six COVID-designated intensive care units (ICUs) at three Emory Healthcare acute-care hospitals in Atlanta, Georgia from March 6, 2020 through April 17, 2020. COVID-19 status was based upon a positive SARS-CoV-2 PCR assay, performed either by the Georgia Department of Public Health, a referral laboratory, or the hospital-based clinical laboratory. Patient data, including sociodemographic information, clinical data, and laboratory data, were obtained from the electronic medical record.

Data were abstracted through April 21, 2020. Data were analyzed using a chi-square or Wilcoxon rank sum test for categorical and continuous variables, respectively, with a 2-sided p-value < 0.05 considered statistically significant (Stata Version 12.1). This study was approved by the Emory University Institutional Review Board.

## Results

### Patient Characteristics

From March 6 through April 17, 2020, 217 critically ill adults with COVID-19 infection were admitted to the ICU (Figure 1A). The median patient age was 64 (interquartile range [IQR] 54–73), with 49 (22.6%) patients who were 75 years or older (Table 1). There were 98 (45.2%) females and the majority of patients were black (153 [70.5%]). Hypertension was the most common comorbid condition (134 [61.7%]), followed by diabetes (99, [45.6%]). Twenty-one patients (9.7%) had morbid obesity, with a body mass index (BMI) of 40 or greater.

**Table 1.** Sociodemographic and baseline clinical characteristics of patients admitted to a COVID-ICU.

| **Characteristic**<br>(n[%] unless otherwise indicated) | **All**<br>**(n=217)** | **Survived ICU**<br>**(n=129)** | **Died in ICU**<br>**(n=52)** | **p-value\*** |
| --- | --- | --- | --- | --- |
| Age, median (IQR) | 64 (54 – 73) | 61 (51 – 69) | 70 (63 – 77) | **< 0.001** |
| Age <55 | 55 (25.3) | 38 (29.5) | 7 (13.5) | **0.024** |
| Age 55-64 | 56 (25.8) | 40 (31.0) | 9 (17.3) | 0.061 |
| Age 65-74 | 57 (26.3) | 31 (24.0) | 18 (34.6) | 0.147 |
| Age ≥ 75 | 49 (22.6) | 20 (15.5) | 18 (34.6) | **0.004** |
| Female | 98 (45.2) | 58 (45.0) | 28 (53.9) | 0.279 |
| Race |  |  |  |  |
| White | 39 (18.0) | 29 (22.5) | 6 (11.5) | 0.092 |
| Black | 153 (70.5) | 90 (69.8) | 38 (73.1) | 0.658 |
| Asian | 7 (3.2) | 1 (0.8) | 2 (3.9) | 0.143 |
| Other/unknown, | 18 (8.3) | 9 (7.0) | 6 (11.5) | 0.314 |
| BMI, median (IQR) | 30 (26 – 35) | 31 (27 – 36) | 29 (26 – 32) | 0.0628 |
| BMI ≥ 40 | 21 (9.7) | 16 (12.4) | 1 (1.9) | **0.029** |
| HTN | 134 (61.7) | 83 (64.3) | 34 (65.4) | 0.894 |
| CHF | 41 (18.9) | 25 (19.4) | 12 (23.1) | 0.577 |
| CAD | 31 (14.3) | 15 (11.6) | 13 (25.0) | **0.024** |
| DM | 99 (45.6) | 60 (46.5) | 26 (50.0) | 0.671 |
| CKD/ESRD | 58 (26.7) | 33 (25.6) | 20 (38.5) | 0.085 |
| Asthma | 19 (8.8) | 13 (10.1) | 3 (5.8) | 0.356 |
| COPD | 21 (9.7) | 14 (10.9) | 4 (7.7) | 0.520 |

\*p-value: chi-square or Wilcoxon rank sum test comparing those who survived vs. died in the ICU.
*BMI = body mass index; CAD = coronary artery disease; CHF = congestive heart failure; CKD = chronic kidney disease; COPD = chronic obstructive pulmonary disease; DM = diabetes mellitus; ESRD = end-stage renal disease; HTN = hypertension; IQR = interquartile range.*

**Figure 1A.**
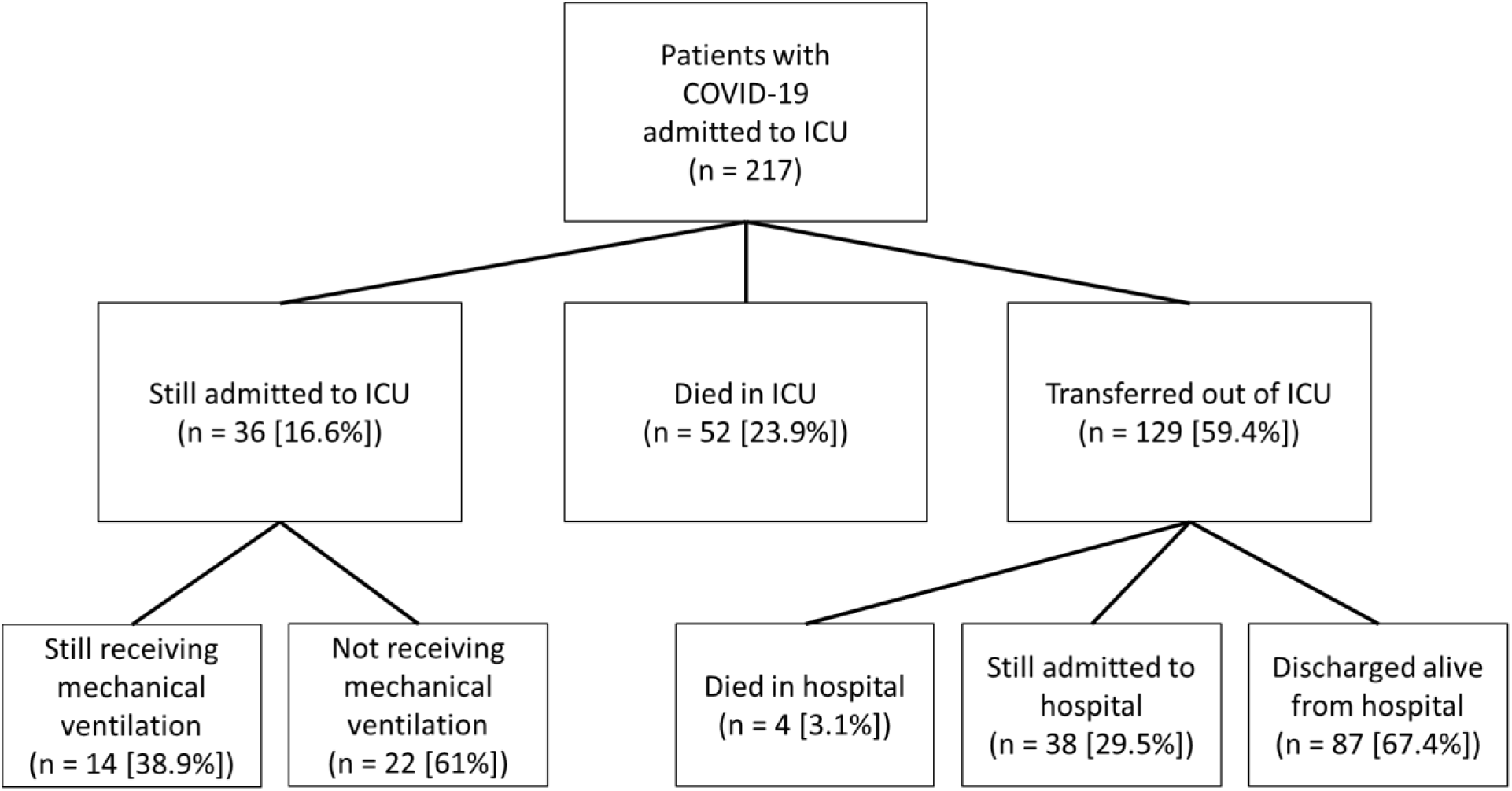
Flow diagram for patients admitted to a COVID-ICU.

**Figure 1B.**
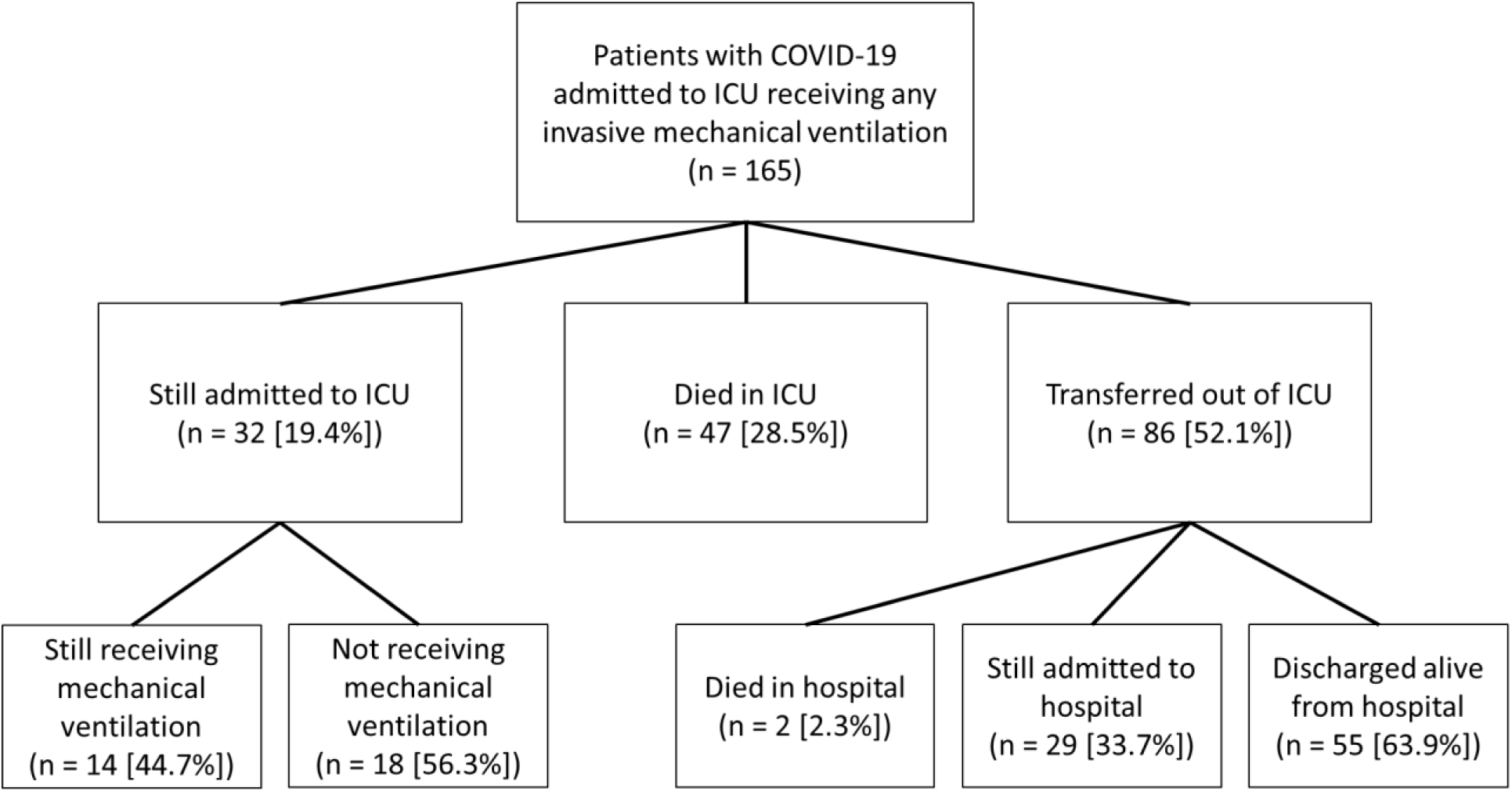
Flow diagram for patients admitted to a COVID-ICU who received any invasive mechanical ventilation

### ICU admission and interventions

Initial ICU clinical findings, critical care interventions, and outcomes are summarized in Table 2. On admission to the ICU, the median SOFA score was 7 (IQR 5–11), the median D-dimer was 1731 ng/ml (IQR 934–6948; upper limit of normal 298 ng/ml), and the median C-reactive protein was 190 mg/L (IQR 126–262; upper limit of normal 10 mg/L). The median initial PaO_2_/FiO_2_ ratio was 132 (IQR 100–178).

**Table 2.** Patient characteristics on ICU admission, ICU clinical interventions, and outcomes.

| **Characteristic**<br>(n[%] unless otherwise indicated) | **All**<br>**(n=217)** | **Survived ICU**<br>**(n=129)** | **Died in ICU**<br>**(n=52)** | **p-value\*** |
| --- | --- | --- | --- | --- |
| **Initial ICU clinical characteristics** |  |  |  |  |
| SOFA, median (IQR) | 7 (5 – 11) | 7 (4 – 10) | 9 (6 – 13) | **0.002** |
| D-dimer, median (IQR) | 1731<br>(934 – 6948) | 1545<br>(890 – 3514) | 3780<br>(1425 – 19468) | **0.002** |
| C-reactive protein, median (IQR) | 190 (126 – 262) | 167 (112 – 241) | 224 (164 – 309) | **0.003** |
| P/F ratio, median (IQR) | 132 (100 – 178) | 144 (107 – 196) | 114 (88 – 148) | **0.001** |
| **ICU Interventions** |  |  |  |  |
| Any Mechanical Ventilation | 165 (76.0) | 86 (66.7) | 47 (90.4) | **< 0.001** |
| Initial static compliance, median (IQR) | 34 (28 – 46) | 33 (28 – 41) | 35 (27 – 47) | 0.778 |
| Ventilator days, median (IQR) | 8 (4 – 12) | 7 (4 – 11) | 8 (4 – 12) | 0.989 |
| Any Vasopressors | 143 (65.9) | 65 (50.4) | 48 (92.3) | **< 0.001** |
| Vasopressor days, median (IQR) | 5 (3 – 9) | 4 (2 – 7) | 6 (3 – 9) | **0.021** |
| Any CRRT/HD | 63 (29.0) | 19 (14.7) | 28 (53.9) | **< 0.001** |
| CRRT/HD days, median (IQR) | 9 (4 – 14) | 11 (6 – 17) | 6 (3 – 12) | **0.033** |
| Inhaled vasodilator | 22 (10.1) | 6 (4.7) | 11 (21.1) | **<0.001** |
| Any ECMO | 4 (1.8) | 2 (1.5) | 0 | 0.367 |
| Hydroxychloroquine | 114 (52.5) | 69 (53.5) | 27 (51.9) | 0.849 |
| ACTT (remdesivir or placebo) | 49 (22.6) | 29 (22.5) | 11 (21.1) | 0.846 |
| **Outcomes** |  |  |  |  |
| ICU days, median (IQR) | 8 (4 – 13) | 7 (3 – 12) | 8 (5 – 12) | 0.382 |
| Hospital days, median (IQR) | 13 (9 – 20) | 15 (9 – 21) | 11 (7 – 14) | **0.0003** |

\*p-value: chi-square or Wilcoxon rank sum test comparing those who survived vs. died in the ICU.
ACTT = adaptive clinical treatment trial; CRRT = continuous renal replacement therapy; ECMO = extracorporeal membrane oxygenation; HD = hemodialysis; IQR = interquartile range; LOS = length of stay; SOFA = Sequential Organ Failure Assessment.

There were 165 (76.0%) patients who received invasive mechanical ventilation. The median static lung compliance on the first day of intubation was 34 ml/cmH_2_O (IQR 28–46). A total of 143 (65.9%) patients required vasopressor support for shock and 63 (29.0%) required renal replacement therapy, either in the form of continuous renal replacement therapy or intermittent hemodialysis. Use of inhaled pulmonary vasodilators was relatively uncommon (22 [10.1%]) and four (1.8%) patients received extracorporeal membrane oxygenation (ECMO). Overall, 114 (52.5%) patients received at least one dose of hydroxychloroquine and 49 (22.6%) patients were enrolled in the NIH-funded adaptive clinical treatment trial (ACTT) of remdesivir (ClinicalTrials.gov NCT04280705).

### ICU Outcomes

Among the 217 patients in the cohort, 129 (59.4%) have been transferred alive from the ICU, 52 (23.9%) died in the ICU (four additional deaths occurred among patients transferred to the floor), and 36 (16.6%) remain in the ICU, of whom 14 (38.9%) are still mechanically ventilated (Figure 1a). The median ICU length of stay for patients still in the ICU is 12 days (IQR 10–18). Among patients who received invasive mechanical ventilation, ICU mortality is 28.5% (47/165) and hospital mortality is 29.7% (49/165) (Figure 1b).

The median age of patients who died was significantly older than for those who survived (70 years [IQR 63–77] vs. 61 years [IQR 51 – 69]; p-value < 0.001) (Table 1). Race and female sex did not differ according to survival, but patients who died were less likely to be morbidly obese and more likely to have coronary artery disease.

Patients who died had a higher Sequential Organ Failure Assessment (SOFA) score on ICU admission (median 9 [IQR 6–13]) than those who survived (median 7 [IQR 4–10]; p-value 0.002) (Table 2). Median D-dimer values were more than two times higher in those who died than those who survived (3780 [IQR 1425 – 19468] vs. 1545 [IQR 890–3514]; p-value 0.002). Similarly, the initial C-reactive protein and PaO_2_/FiO_2_ ratios were significantly worse in patients who died in the ICU (p-values 0.003 and 0.001, respectively). Compared to those who survived, patients who died in the ICU were more likely to have respiratory failure requiring invasive mechanical ventilation (90.4% vs. 66.7%; p-value < 0.001), shock requiring vasopressors (92.3% vs. 50.4%, p-value <0.001), and renal failure requiring renal replacement therapy (53.9% vs 14.7%, p value <0.001).

Overall, mortality among patients who received vasopressors was 33.6% (48/143), while 28 (44.5%) of 63 patients with renal failure requiring renal replacement therapy died. Half of the patients who received inhaled vasodilator for refractory hypoxemia died (11/22). Among the 52 patients who died in the ICU, including three patients who had an advance directive not to be intubated, the median time from ICU admission to death was 8 days (IQR 5–12).

## Discussion

Our early experience with this large cohort of critically ill patients with COVID-19 demonstrates a mortality rate of 25.8% overall, which is substantially lower than the 50 – 97% reported in the published literature to date.^1-6^ Additionally, the 29.7% mortality for the approximately three-quarters of patients in our cohort who required mechanical ventilation is also markedly lower than previous reports. These data indicate that a majority of critically ill patients with COVID-19 can have good clinical outcomes and support the ongoing use of mechanical ventilation for patients with acute respiratory failure.

In some of the earliest reports of COVID-19 from Wuhan, mortality rates among those admitted to ICUs ranged from 52–62%, and increased to 86–97% among those requiring invasive mechanical ventilation.^5,6,15,16^ In more recent data from the United Kingdom, 67% of those who had received mechanical ventilation died, as compared to 22% of patients intubated with viral pneumonia in the preceding three years.^3^ Early reports of smaller cohorts from Seattle, where some of the first COVID-19 outbreaks occurred in the United States, indicated that 50–67% of patients admitted to the ICU and 71– 75% of those receiving invasive mechanical ventilation died.^1,2^ A recently published report from New York also found high mortality of 88.1% among those who required mechanical ventilation.^4^

Taken together, these reports have raised concerns that survival among those receiving mechanical ventilation is exceedingly poor.^11-14,17^ In contrast to the majority of prior reports, our data provide evidence that mortality rates in COVID-19 can be comparable to those seen with ARDS and other infectious pneumonias.^7-10^

In our cohort, mortality was associated with older age, with 30.1% mortality in those age 65 and above as compared to 9.1% in those under age 55. Mortality was also associated with the presence of coronary artery disease, severity of illness on arrival to the ICU, and need for ICU interventions including mechanical ventilation, vasopressor support, renal replacement therapy and inhaled vasodilators. Approximately half of our cohort received at least one dose of hydroxychloroquine and nearly one-quarter received at least one dose of a study drug as part of the ACTT trial (remdesivir vs. placebo), but there was no difference in survival for either of those groups. Finally, although 36 (16.6%) patients are still admitted to the ICU at the time of this report, 87 (40.1%) have been discharged from the hospital.

Several local and regional considerations may have influenced the observed outcomes. First, the arrival and peak of the COVID-19 pandemic in Georgia were later than in many of the regions from earlier reports. This delay provided time to establish organizational structures, acquire equipment, prepare personnel, create consensus-driven clinical protocols, and align resources across a large healthcare system. In addition, while patient volumes did merit the re-designation of several specialty intensive care units as COVID ICUs, all critically ill patients with COVID-19 were admitted to pre-existing ICUs and cared for by critical care teams with experience managing acute respiratory failure and at standard patient-to-provider ratios. Further, while an analysis of the impact of clinical interventions on survival is beyond the scope of this brief report, our internal guidelines emphasized early intubation and standard lung-protective ventilation strategies. Future studies are needed to better understand the impact of individual patient risk factors, clinical interventions and treatments, and health system factors that can improve mortality in the face of this global pandemic.

## Conclusion

In a cohort of critically ill adults with COVID-19, we report an early mortality rate of 25.8% overall and 29.7% for patients who received mechanical ventilation. While there may be a several factors underlying these findings, these results suggest that most patients with acute respiratory failure from COVID-19 may recover, even with severe disease requiring intubation and mechanical ventilation.

## Data Availability

Investigators may contact the authors regarding the availability of data.

## Conflict of interest disclosures

None reported.

## Funding

This work was supported by the following grants: NIH/NIAID K23 AI134182 (SCA), NIH/CTSA UL1TR002378. The content is solely the responsibility of the authors and does not necessarily represent the official views of the National Institutes of Health.

## Role of the Funder/Sponsor

The funders had no role in the design and conduct of the study; collection, management, analysis, and interpretation of the data; preparation, review, or approval of the manuscript; and decision to submit the manuscript for publication.

## Acknowledgements

We would like to extend our most profound thanks and gratitude to our colleagues in the Emory Critical Care Center and Emory Healthcare who have worked so hard to provide excellent clinical care during this global pandemic.

Emory COVID-19 Quality and Clinical Research Collaborative Members (in alphabetical order): Max W. Adelman, Scott Arno, Sara C. Auld, Theresa Barnes, William Bender, James M. Blum, Gaurav Budharani, Stephanie Busby, Laurence Busse, Mark Caridi-Scheible, David Carpenter, Nikulkumar Chaudhari, Craig M. Coopersmith, Lisa Daniels, Jane Fazio, Babar Fiza, Eliana Gonzalez, Ria Gripaldo, Charles Grodzin, Robert Groff, Alfonso C. Hernandez-Romieu, Max Hockstein, Dan Hunt, Craig S. Jabaley, Jesse T. Jacob, Colleen Kraft, Greg S. Martin, Samer Melham, Nirja Mehta, Chelsea Modlin, David J. Murphy, Mia Park, Deepa Patel, Cindy Powell, Amit Prabhaker, Jeeyon Rim, Ramzy Rimawi, Chad Robichaux, Nicholas Scanlon, Milad Sharifpour, Bashar Staitieh, Michael Sterling, Jonathan Suarez, Colin Swenson, Nancy Thakkar, Alexander Truong, Hima Veeramachaneni, Alvaro Velasquez, Michael Waldmann, Max Weinmann, Thanushi Wynn, and Joel Zivot.

## Notes

### Competing Interest Statement

The authors have declared no competing interest.

